# Modelled health impacts of three stakeholder-selected policies to support healthy and environmentally sustainable population diets

**DOI:** 10.1101/2025.08.03.25332921

**Authors:** Christine Cleghorn, Christina McKerchar, Bruce Kidd, Cliona Ni Mhurchu

## Abstract

Food production and consumption impacts both planetary and population health. This research aimed to estimate the impacts of three stakeholder-selected food policies for the New Zealand (NZ) population. These were modelled using a proportional multistate lifetable model through to health gains, health equity impacts, and health system costs/cost savings. Exempting core, sustainable foods from Goods and Services Tax resulted in 87,340 health adjusted life years (HALYs) (95% Uncertainty Interval (UI): 72,590 to 104,140) and health system cost savings of $925.0 Million (M), UI: $661.5 to $1220.3M. Education about healthy sustainable foods generated small health impacts (1,170 HALYs, UI: 930 to 1,430) and health system cost savings ($9.6M, UI: $6.8 to $12.9M). Māra kai (gardening for food) and community gardens generated 8,040 HALYs (UI: 4,720 to 11,800) and would cost the government $364.3M (UI: $267.6 to $464.6M). All scenarios produced more age-standardised per capita health gain for Māori (Indigenous population) than non-Māori. The first two policies were cost saving while the māra kai and community gardens policy was borderline cost-effective. These results show that these policies have the potential to improve population health and reduce NZ ethnic health inequities. Due to the up-front investment needed for these policies and the lag in generating health system costs savings, research into the policies wider benefits need to be considered alongside these results. Results could be used to inform diet policy in high-income countries. For NZ, careful policy design involving relevant stakeholders, including Māori communities, is needed to ensure health benefits are realised.

**Highlights:**

- Uses rich qualitative data on stakeholder perspectives to inform policy design
- Presents three food policies of varying designs to inform real world policy
- Identifies cost saving policies which could potentially reduce health inequities
- Policies require large up-front costs or loss of governmental income
- Health system cost savings are expected in the medium to long term

## 1. Introduction

Dietary intake is an important risk factor for cardiovascular disease, diabetes and various cancers (Afshin, Sur et al. 2019), with an appreciable reduction in disease burden expected should population diets shift towards healthier options. High intakes of sodium and low intakes of wholegrains and fruits are the leading dietary risk factors for death and disability adjusted life-years (Afshin, Sur et al. 2019) with obesity due to poor diet leading to further incidence of chronic disease. NZ diets are low in fruit and vegetables and contribute to high rates of overweight and obesity (University of Otago and Ministry of Health 2011, Ministry of Health 2023).

NZ has a history of colonisation, resulting in health inequities experienced by Māori, the Indigenous population (Reid and Robson 2000). This has impacted access to healthy foods for Māori. Māori are less likely to eat recommended servings of fresh fruit and vegetables, and are more likely to eat processed meat than the general population, dietary practices which are risk factors for non-communicable diseases (Afshin, Sur et al. 2019). These inequities could be reduced through policy change focused on ensuring equity of health status for Māori, working in partnership with Māori communities as guided by Te Tiriti o Waitangi (The Treaty of Waitangi, glossary of Māori words included in Supplementary material)(Waitangi Tribunal 2019, Public Health Advisory Committee 2024).

Alongside the health consequences directly associated with poor diet, the health effects of environmental degradation are widely apparent and could pose extreme risks to human health through disruptive climate change and other environmental impacts (Corvalan, Hales et al. 2005, Whitmee, Haines et al. 2015). A recent Lancet Commission described the interrelated combination of obesity, undernutrition and climate change as a global syndemic (Swinburn, Kraak et al. 2019). The global food system is responsible for up to 29% of all anthropogenic GHG emissions (Vermeulen, Campbell et al. 2012) with agriculture being responsible for half of NZ’s GHG emissions (Ministry for the Environment 2021). Shifts in diet away from animal protein and processed foods and towards more vegetables, legumes, wholegrains and fruit are likely to have health and environmental co-benefits (Springmann, Godfray et al. 2016).

The EAT-Lancet Commission proposed a ‘Great Food Transformation’ including a global reference diet to bring the six food system related earth system processes into safe operating boundaries (Willett, Rockström et al. 2019). The reference diet is high in vegetables, legumes, wholegrains, fruit and plant sources of protein and low in animal sources of protein. The Commission takes a global perspective and notes that their framework does not provide a plan for translating these global targets to national targets or practical solutions to meet these targets. There exists, therefore, an important research gap on how individual countries can shift current population diets towards this healthier and more sustainable reference diet.

Figure 1 illustrates the food system context in NZ, alongside the potential health and environmental benefits of shifts towards sustainable dietary intakes. Results from stakeholder consultation undertaken within the current project (Kidd B, Enright H et al. 2025) outlined the importance of strong local food networks and being able to grow and collect food locally for New Zealanders, all of which impact on food security and food system resilience. This local food system sits within a wider national food system driven by export markets which can overshadow the importance of providing healthy, affordable and sustainable food for New Zealanders.

**Figure 1.**
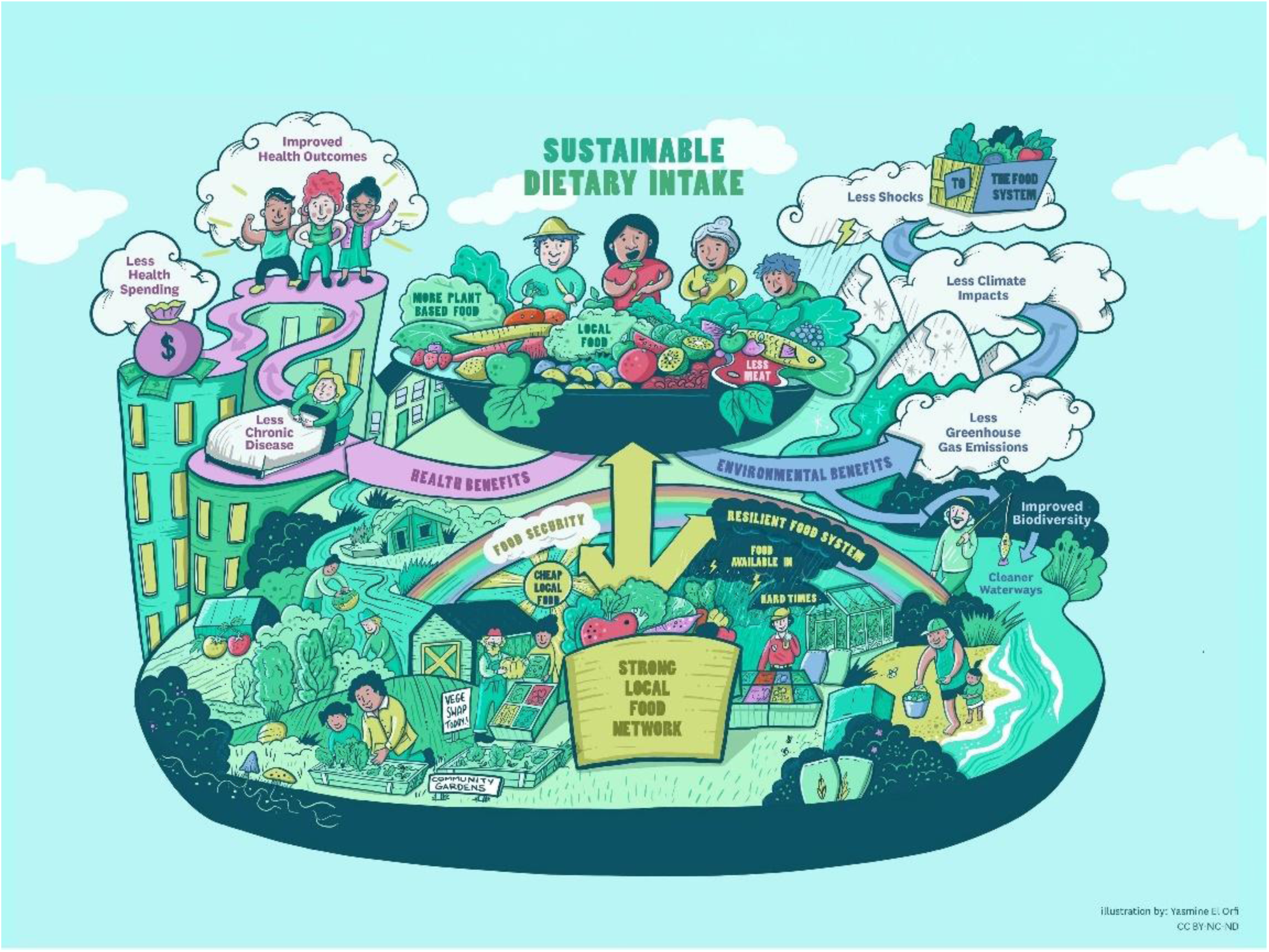
Health, environmental and resilience co-benefits of a sustainable food system supporting sustainable dietary intake.

Research on the cost-effectiveness and scope of health gain for policies designed to support healthy sustainable consumption in NZ can help policy makers select appropriate strategies to implement. This article aims to estimate the health, health system costs, and equity impacts of three stakeholder identified dietary policies to support New Zealanders to consume more healthy and sustainable foods.

## 2. Methods

### 2.1 Selection of modelled policies

Modelled policies were selected through a 3-stage process using qualitative interviews and focus groups with key stakeholders (Kidd B, Enright H et al. 2025). Six focus groups with a total of 57 stakeholders from the Ministry of Health, Ministry for the Environment, Ministry for Primary Industries, a civil society group recruited through community organisations and flyers and contributors from two Māori communities, one based in an urban area and one based more rurally. Interviews were also carried out with 13 academic and food industry representatives. Stakeholders initially suggested a range of policies that could potentially help support New Zealanders to consume more healthy and environmentally sustainable food (N=111). The policies that had direct evidence on the effect on dietary intake (specifically the dietary risk factors we model (Cleghorn, Blakely et al. 2017)) were taken to the subsequent stage (N=14). In this stage stakeholders and project advisors rated the policies on five criteria (improve Māori health, improve NZ diets, improve the environment, ease of implementation, acceptability). Average ratings were considered in the whole group and in Māori participants specifically. Top rated policies were taken through to the final stakeholder consultation. Here stakeholders discussed how best to implement these policies in the NZ context. The design of the five modelled policies were based on the details of these discussions. In this paper we present the three policies modelled in the whole population, excluding two aimed at children only. The proportion of the population that are affected by these policies are presented in Figure 2 and Table 1. The costs associated with implementing these policies are presented in Figure 3 and Table 1.

**Figure 2.**
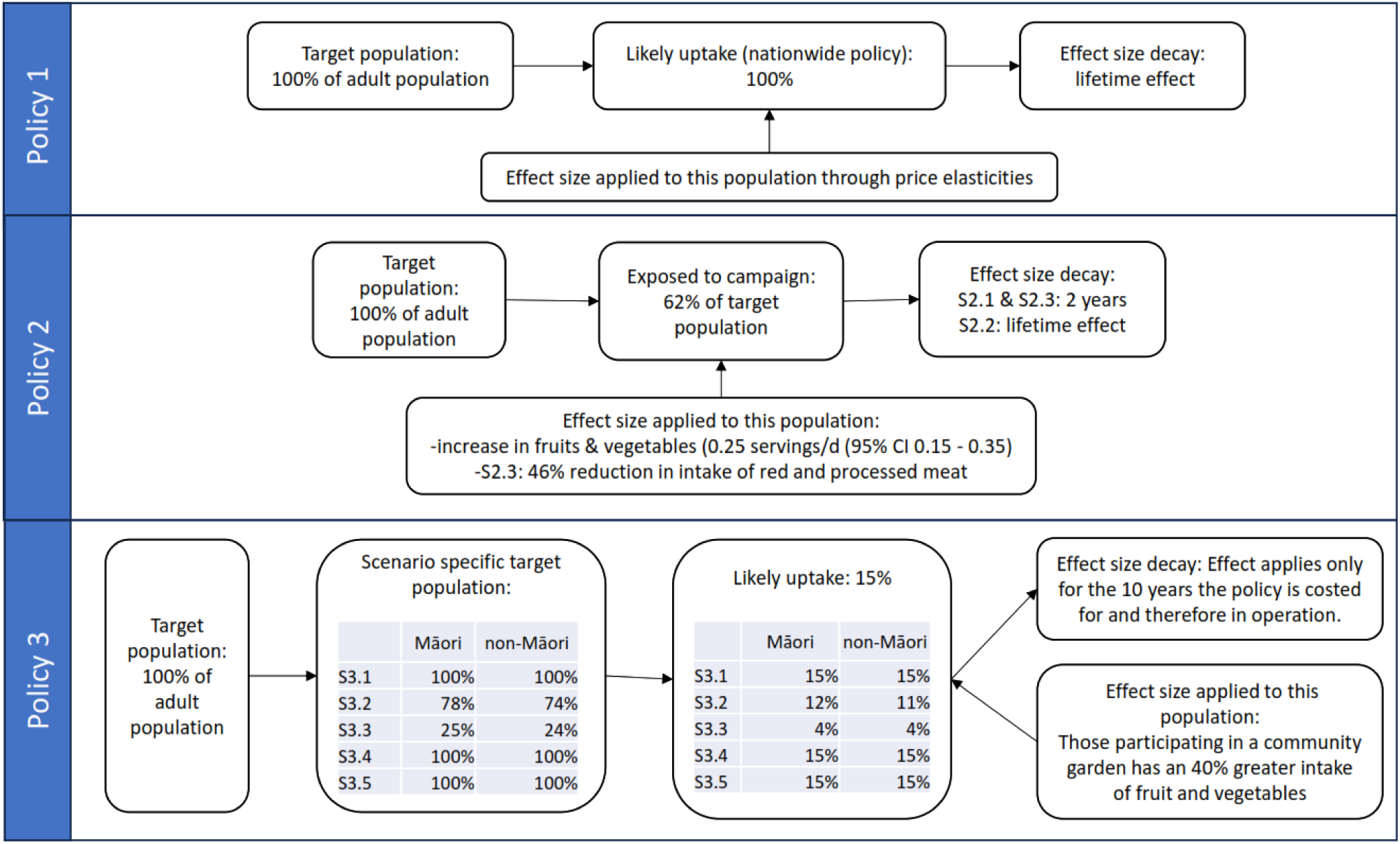
Proportion of the population affected by all policies (S2.1: Effect size for 2 years, S2.2: Effect size for lifetime of cohort, S2.3: S2.1 plus effect on red and processed meat intake, S3.1: All marae and suburbs in NZ (0.5 FTE/garden), S3.2: All marae and matched number of suburbs in NZ (0.5 FTE/garden), S3.3: 1/3 of Marae and matched number of suburbs in NZ (0.5 FTE/garden), S3.4: All marae and suburbs in NZ (1 FTE/garden), S3.5: All marae and suburbs in NZ (1 FTE/15 garden)).

**Figure 3.**
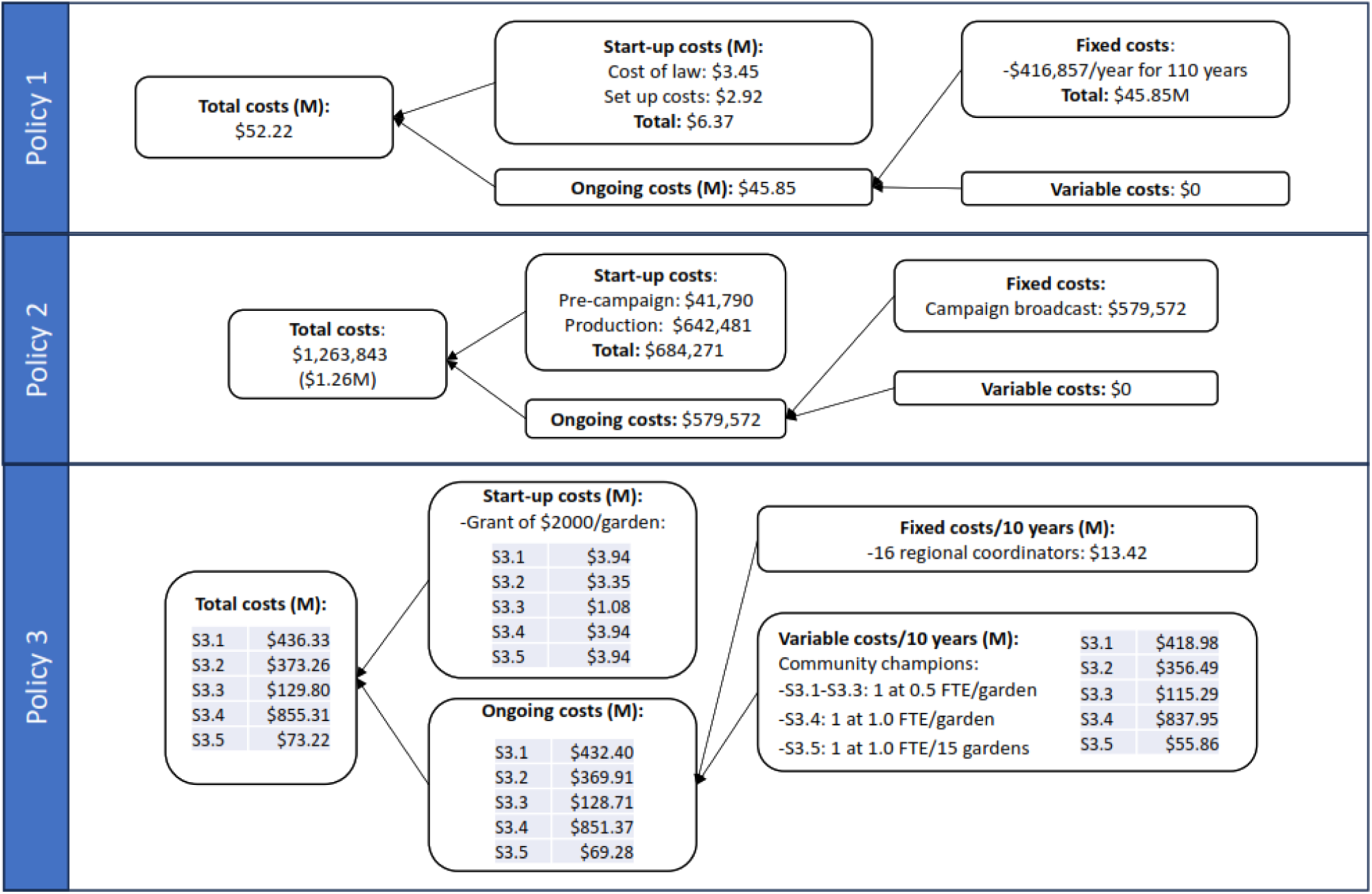
Costs of implementing all policies (S2.1: Effect size for 2 years, S2.2: Effect size for lifetime of cohort, S2.3: S2.1 plus effect on red and processed meat intake, S3.1: All marae and suburbs in NZ (0.5 FTE/garden), S3.2: All marae and matched number of suburbs in NZ (0.5 FTE/garden), S3.3: 1/3 of Marae and matched number of suburbs in NZ (0.5 FTE/garden), S3.4: All marae and suburbs in NZ (1 FTE/garden), S3.5: All marae and suburbs in NZ (1 FTE/15 garden)). M: Million.

**Table 1.**
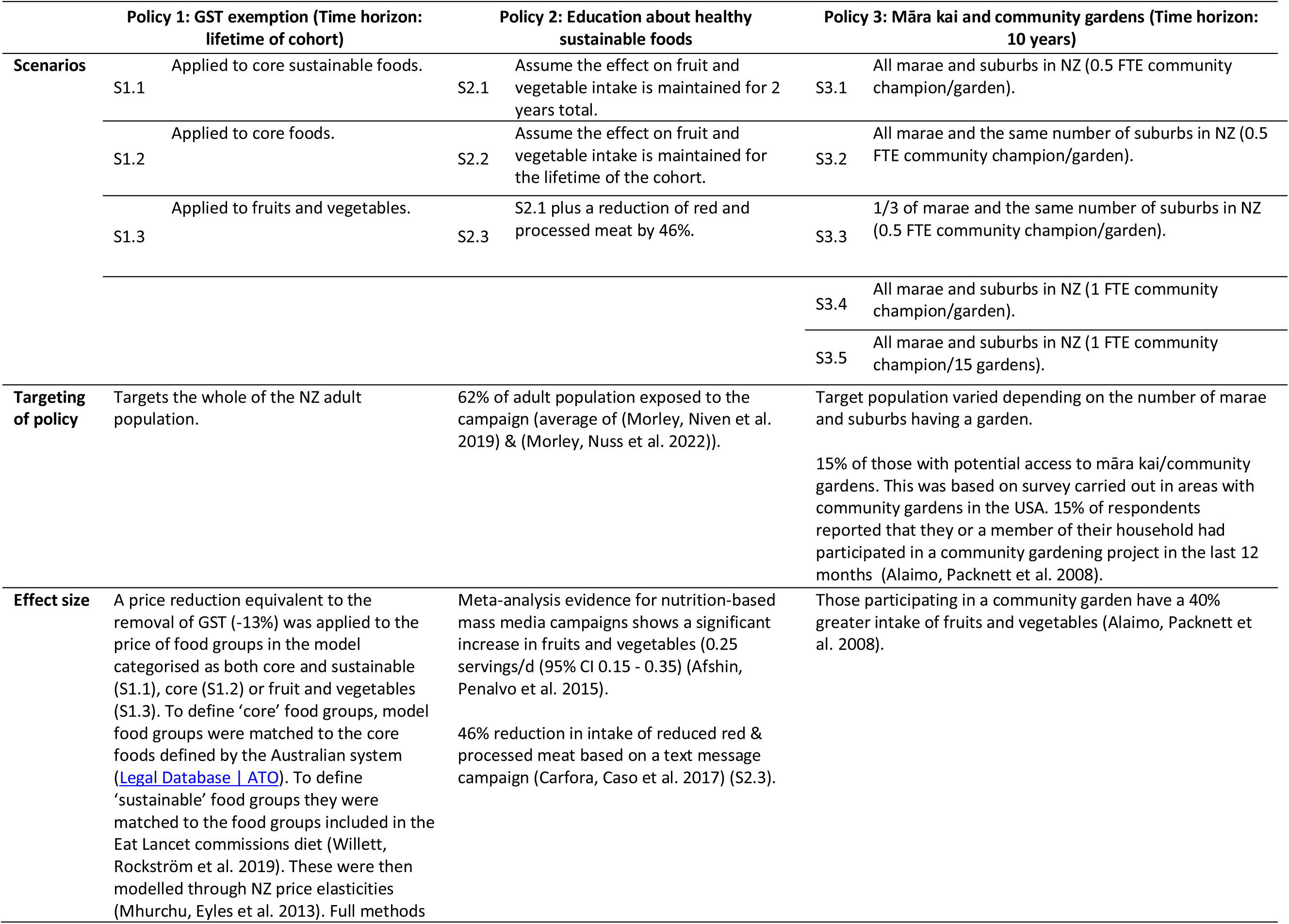
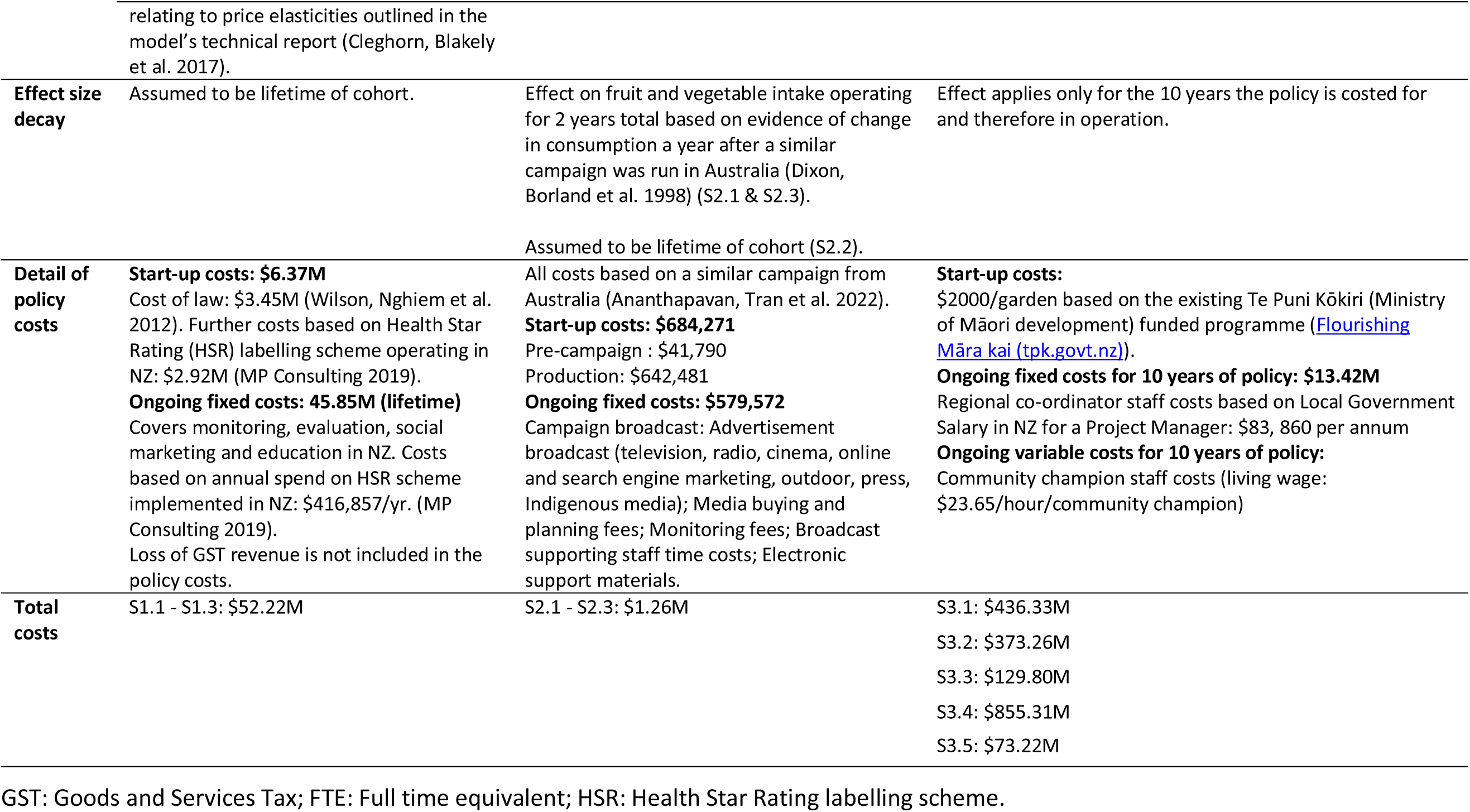
Intervention parameters for the three policies.

|  | **Policy 1: GST exemption (Time horizon: lifetime of cohort)** | **Policy 2: Education about healthy sustainable foods** | **Policy 3: Māra kai and community gardens (Time horizon: 10 years)** |
| --- | --- | --- | --- |
| **Scenarios** | Applied to core sustainable foods.<br>S1.1 | S2.1 Assume the effect on fruit and vegetable intake is maintained for 2 years total. | S3.1 All marae and suburbs in NZ (0.5 FTE community champion/garden). |
|  | Applied to core foods.<br>S1.2 | S2.2 Assume the effect on fruit and vegetable intake is maintained for the lifetime of the cohort. | S3.2 All marae and the same number of suburbs in NZ (0.5 FTE community champion/garden). |
|  | Applied to fruits and vegetables.<br>S1.3 | S2.3 S2.1 plus a reduction of red and processed meat by 46%. | S3.3 1/3 of marae and the same number of suburbs in NZ (0.5 FTE community champion/garden). |
|  |  |  | S3.4 All marae and suburbs in NZ (1 FTE community champion/garden). |
|  |  |  | S3.5 All marae and suburbs in NZ (1 FTE community champion/15 gardens). |
| **Targeting of policy** | Targets the whole of the NZ adult population. | 62% of adult population exposed to the campaign (average of (Morley, Niven et al. 2019) & (Morley, Nuss et al. 2022)). | Target population varied depending on the number of marae and suburbs having a garden.<br>15% of those with potential access to māra kai/community gardens. This was based on survey carried out in areas with community gardens in the USA. 15% of respondents reported that they or a member of their household had participated in a community gardening project in the last 12 months (Alaimo, Packnett et al. 2008). |
| **Effect size** | A price reduction equivalent to the removal of GST (-13%) was applied to the price of food groups in the model categorised as both core and sustainable (S1.1), core (S1.2) or fruit and vegetables (S1.3). To define 'core' food groups, model food groups were matched to the core foods defined by the Australian system (Legal Database ATO). To define 'sustainable' food groups they were matched to the food groups included in the Eat Lancet commissions diet (Willett, Rockström et al. 2019). These were then modelled through NZ price elasticities (Mhurchu, Eyles et al. 2013). Full methods relating to price elasticities outlined in the model's technical report (Cleghorn, Blakely et al. 2017). | Meta-analysis evidence for nutrition-based mass media campaigns shows a significant increase in fruits and vegetables (0.25 servings/d (95% CI 0.15 - 0.35) (Afshin, Penalvo et al. 2015).<br>46% reduction in intake of reduced red & processed meat based on a text message campaign (Carfora, Caso et al. 2017) (S2.3). | Those participating in a community garden have a 40% greater intake of fruits and vegetables (Alaimo, Packnett et al. 2008). |
| **Effect size decay** | Assumed to be lifetime of cohort. | Effect on fruit and vegetable intake operating for 2 years total based on evidence of change in consumption a year after a similar campaign was run in Australia (Dixon, Borland et al. 1998) (S2.1 & S2.3). | Effect applies only for the 10 years the policy is costed for and therefore in operation. |
|  |  | Assumed to be lifetime of cohort (S2.2). |  |
| **Detail of policy costs** | **Start-up costs: \$6.37M**<br>Cost of law: \$3.45M (Wilson, Nghiem et al. 2012). Further costs based on Health Star Rating (HSR) labelling scheme operating in NZ: \$2.92M (MP Consulting 2019).<br>**Ongoing fixed costs: 45.85M (lifetime)**<br>Covers monitoring, evaluation, social marketing and education in NZ. Costs based on annual spend on HSR scheme implemented in NZ: \$416,857/yr. (MP Consulting 2019).<br>Loss of GST revenue is not included in the policy costs. | All costs based on a similar campaign from Australia (Ananthapavan, Tran et al. 2022).<br>**Start-up costs: \$684,271**<br>Pre-campaign : \$41,790<br>Production: \$642,481<br>**Ongoing fixed costs: \$579,572**<br>Campaign broadcast: Advertisement broadcast (television, radio, cinema, online and search engine marketing, outdoor, press, Indigenous media); Media buying and planning fees; Monitoring fees; Broadcast supporting staff time costs; Electronic support materials. | **Start-up costs:**<br>\$2000/garden based on the existing Te Puni Kōkiri (Ministry of Māori development) funded programme ([Flourishing Māra kai (tpk.govt.nz)](https://www.flourishingmāorikai.govt.nz/)).<br>**Ongoing fixed costs for 10 years of policy: \$13.42M**<br>Regional co-ordinator staff costs based on Local Government Salary in NZ for a Project Manager: \$83, 860 per annum<br>**Ongoing variable costs for 10 years of policy:**<br>Community champion staff costs (living wage: \$23.65/hour/community champion) |
| **Total costs** | S1.1 - S1.3: \$52.22M | S2.1 - S2.3: \$1.26M | S3.1: \$436.33M<br>S3.2: \$373.26M<br>S3.3: \$129.80M<br>S3.4: \$855.31M<br>S3.5: \$73.22M |

GST: Goods and Services Tax; FTE: Full time equivalent; HSR: Health Star Rating labelling scheme.

### 2.2 Policy 1: Goods and Services Tax (GST) exemption

Under the current GST system in NZ, almost all goods and services are taxed at the rate of 15%, this includes all sold food. The design of the current NZ system, with it’s single rate and few exemptions, is intended to simplify the tax system and reduce administrative costs. This modelled policy simulated removal of GST from certain foods. Three scenarios were modelled under this policy:

- S1.1: GST exemption applied to core sustainable foods.
- S1.2: GST exemption applied to core foods.
- S1.3: GST exemption applied to fruits and vegetables.

Core foods were defined using the Australian definitions of core foods. To define ‘core’ NZ food groups, model food groups were matched to the core foods defined by the Australian system (<u>Legal</u> <u>Database | ATO</u>). Sustainable foods were defined as those food groups included in the EAT Lancet diet (Willett, Rockström et al. 2019). To be included in S1.1 food groups needed to fit both categories.

In practice, regulation would likely be needed to ensure retailers did not absorb the cost savings from these price reductions and savings would be passed onto consumers.

#### 2.2.1 Population dietary intake data used in Policy 1: GST exemption

Dietary intakes were sourced from 24-h dietary recalls reported in the most recent (2008/09) New Zealand Adult Nutrition Survey (NZANS, n=4721 individuals aged 15 years or older). Baseline dietary intake of 346 food groups was used in modelling GST exemption. Changes in price were modelled through NZ specific price elasticities to estimate the resulting change in dietary intake. This average change in dietary intake was used to calculate the change in dietary risk factors by sex and ethnic (Māori and non-Māori) groups which was subsequently linked through to the diet proportional Multistate Lifetable model (pMSLT).

Initial price elasticities were from the SPEND Study, conducted for NZ (Nnoaham, Sacks et al. 2009, Mhurchu, Eyles et al. 2013). These are in a 24 by 24 matrix (see Figure 2 in the technical report (Cleghorn, Blakely et al. 2017)) of own- and cross-PEs (with standard errors for default uncertainty). These 24 food groups were matched to the 346 food groups used in the intervention model. This gives us 24 overall food groups and 338 food subgroups (ignoring 5 ‘alcoholic beverage’ groups, 2 ‘dietary supplement’ groups and 1 ‘not applicable’ group). The 24 by 24 price elasticity matrix was expanded to a 338 by 338 matrix following the methodology outlined in the technical report (Cleghorn, Blakely et al. 2017).

### 2.3 Policy 2: Education about healthy sustainable foods

This modelled policy was a mass media campaign which would encourage consumption of healthy, sustainable foods in the general population. Such a policy could utilise TV advertising, online, radio and print media. This would (likely) be led by the Ministry of Health (with input from iwi and other relevant government agencies such as Ministry for the Environment and Ministry for Primary Industries). Alongside a focus on healthy eating, it would include key topics such as how Aotearoa NZ grows food, how food production practices affect people and the environment and promoting meat-free meals (e.g. at least once a week). Input to the design of such a mass media campaign by Māori would be essential to ensure this policy does not contribute to dietary and health inequities. Three scenarios were modelled under this policy:

- S2.1: Assume the increase in fruit and vegetable intake is maintained for two years total.
- S2.2: Assume the effect on fruit and vegetable intake is maintained for the lifetime of the cohort.
- S2.3: S2.1 plus a reduction of red and processed meat.

There is a lack of evidence on the long-term effect of mass media campaigns on dietary intake so the first and last scenarios were only modelled to change dietary intake (Afshin, Abioye et al. 2013, Afshin, Penalvo et al. 2015) in the short term (Dixon, Borland et al. 1998). The second scenario modelled a lifelong effect to illustrate the potential of a well-designed long-term mass media campaign. In the final scenario we modelled a 46% reduction in red and processed meat intake for a year of the cohort’s lifetime. However, the evidence on the effect of a mass media campaign on red and processed meat intake is weak: participants were only followed up for 1 week (Carfora, Caso et al. 2017).

### 2.4 Policy 3: Māra kai (food garden informed by mātauranga Māori) and community gardens

This modelled policy would provide land, funding and a community champion to set up and maintain community gardens in suburbs and māra kai at or near marae around the country. For those marae and suburbs that already have māra kai or community gardens provided support could be used to help maintain and expand their gardens. Five scenarios, which varied the number of gardens and the level of support, were modelled under this policy:

- S3.1: Māra kai would be set up in all marae (N=773) and community gardens in all suburbs (N=1044) in NZ. A part-time (0.5 full-time equivalent, FTE) community champion would be paid for each garden.
- S3.2: Māra kai would be set up in all marae (N=773) and an equivalent number of community gardens in some suburbs (N=773) in NZ. A part-time (0.5 FTE) community champion would be paid for each garden.
- S3.3: Māra kai would be set up in a third of all marae (N=250) and an equivalent number of community gardens in some suburbs (N=250) in NZ. A part-time (0.5 FTE) community champion would be paid for each garden.
- S3.4: Māra kai would be set up in all marae (N=773) and community gardens in all suburbs (N=1044) in NZ. A full-time community champion would be paid for each garden.
- S3.5: Māra kai would be set up in all marae (N=773) and community gardens in all suburbs (N=1044) in NZ. A full-time community champion would be paid for, however they will be responsible for supporting 15 māra/gardens per person.

Robust funding to ensure longevity and centring community champions to help run the gardens would be important elements of this policy. The policy would be financed nationally and distributed to relevant local communities. This policy would require high-value land with good soil quality being allocated preferentially to the gardens. The gardens would incorporate food hubs with connections to existing farmers and gardens which would be co-ordinated by the paid community champions.

This policy would aim to build and help sustain existing gardens/māra and would centre community champions for the māra. Key considerations for māra kai include having mana motuhake, security and self-determination at the core, and access to māra marae for urban Māori. Education/knowledge transfers would be included alongside the gardens, such as the importance of gardening, how to cook and eat the food, and using māra kai as a space to tell Māori stories through kai.

### 2.5 Proportional multistate lifetable model

The change in dietary risk factors were modelled through to health adjusted life years (HALYs), health equity between Māori and non-Māori and health system costs or cost savings from diet-related diseases using an established diet pMSLT macrosimulation model (Cleghorn, Blakely et al. 2017, Cleghorn, Wilson et al. 2018).

The NZ population in 2011 (n=4:4 million) were modelled out to death or until age 110 in the diet pMSLT model. The model is parameterized with rich national data by sex, age, and ethnicity. The diet pMSLT model includes a range of dietary risk factors (high intake of red meat, processed meat and sugar-sweetened beverages as well as low intake of fruit, vegetables, and nuts and seeds) and 8 diseases associated with one or more modelled dietary risk factors: coronary heart disease, stroke, type 2 diabetes, and multiple cancers (esophageal, colorectal, ovarian, head and neck, lung).

In this model the entire NZ population that was alive in 2011 is simulated over the remainder of their lives. The model consists of a main lifetable with projected all-cause mortality and morbidity rates and diet-related disease-life tables running in parallel. Disease incidence, case-fatality and remission (cancers only) determine the proportion of the population in each parallel disease state. Changes in dietary risk factors resulting from the policies are combined with disease-specific relative risks obtained from the Global Burden of Disease (GBD) study (Forouzanfar, Alexander et al. 2015) through population impact fractions that alter the incidence of diet-related diseases in the model. The change in incidence of disease is modelled through to a reduction in morbidity and mortality from each disease which are combined to calculate HALYs. Morbidity is quantified for each disease as prevalent years of life lived with disability from the NZ Burden of Disease Study (Ministry of Health 2013), divided by the number in the total population.

Sex- and age-specific health system costs (in NZ dollars, 2011) were calculated from individually-linked data for publicly-funded (and some privately-funded) health events occurring 2006-10 (Kvizhinadze, Nghiem et al. 2016). These included hospitalisation costs, inpatient procedures, outpatient costs, pharmaceutical costs, laboratory costs, and expected primary care usage costs. Costs were calculated for individuals without a diet-related disease and who were not in the last six months of their life (sex- and age-specific), as well as disease-specific excess costs for people in the first year of diagnosis, last six months of life if dying of the given disease, and otherwise prevalent cases of each disease. The modelled intervention costs are outlined in 95% uncertainty intervals (95% UI) are generated by Monte Carlo simulations (n = 2000) to reflect uncertainty around the input parameters in the model. Ethnic inequities in health were quantified through estimating HALYs for Māori and non-Māori separately and presenting the ratio of per capita HALY gains by ethnicity. It was assumed that the targeting of policies (except for policy 3: Māra kai and community gardens which varied by ethnicity), effect size and effect size decay were the same for Māori and non-Māori. Population size, demographics, dietary intake, background mortality and morbidity and diseases rates differed between Māori and non-Māori.

### 2.6 Additional scenario analyses

In the main analyses the Māori population are disadvantaged due to higher background morbidity and mortality rates, i.e. there is a lesser “envelope” for potential health gains. To remove this disadvantage, we present an equity analysis (McLeod, Blakely et al. 2014) where we set Māori background morbidity and mortality rates at the same level as non-Māori values. Each scenario was also run with no discounting so health gain in the future is valued the same as health gain in the present (Supplementary material, supplementary table 1).

## 3 Results

### 3.1 Policy 1: GST exemption

Scenario 1.1, GST exemption on foods that are categorised as both core and sustainable results in modest increases in fruit and vegetable intakes (15g and 28g respectively), with very small decreases in red and processed meat and sugar sweetened beverages (SSBs, Table 2). GST exemption on core foods only (S1.2) followed a similar pattern but with smaller changes in fruit and vegetables, small increases in meat and a larger decrease in SSBs (7g). GST exemption on fruit and vegetables only (S1.3) also resulted in modest increases in fruit and vegetables, 17g and 27g respectively, very small decreases in red and processed meat of 1g each and of SSBs (2g). Nuts and seed intakes did not change more than 0.5g under any scenario.

**Table 2.**
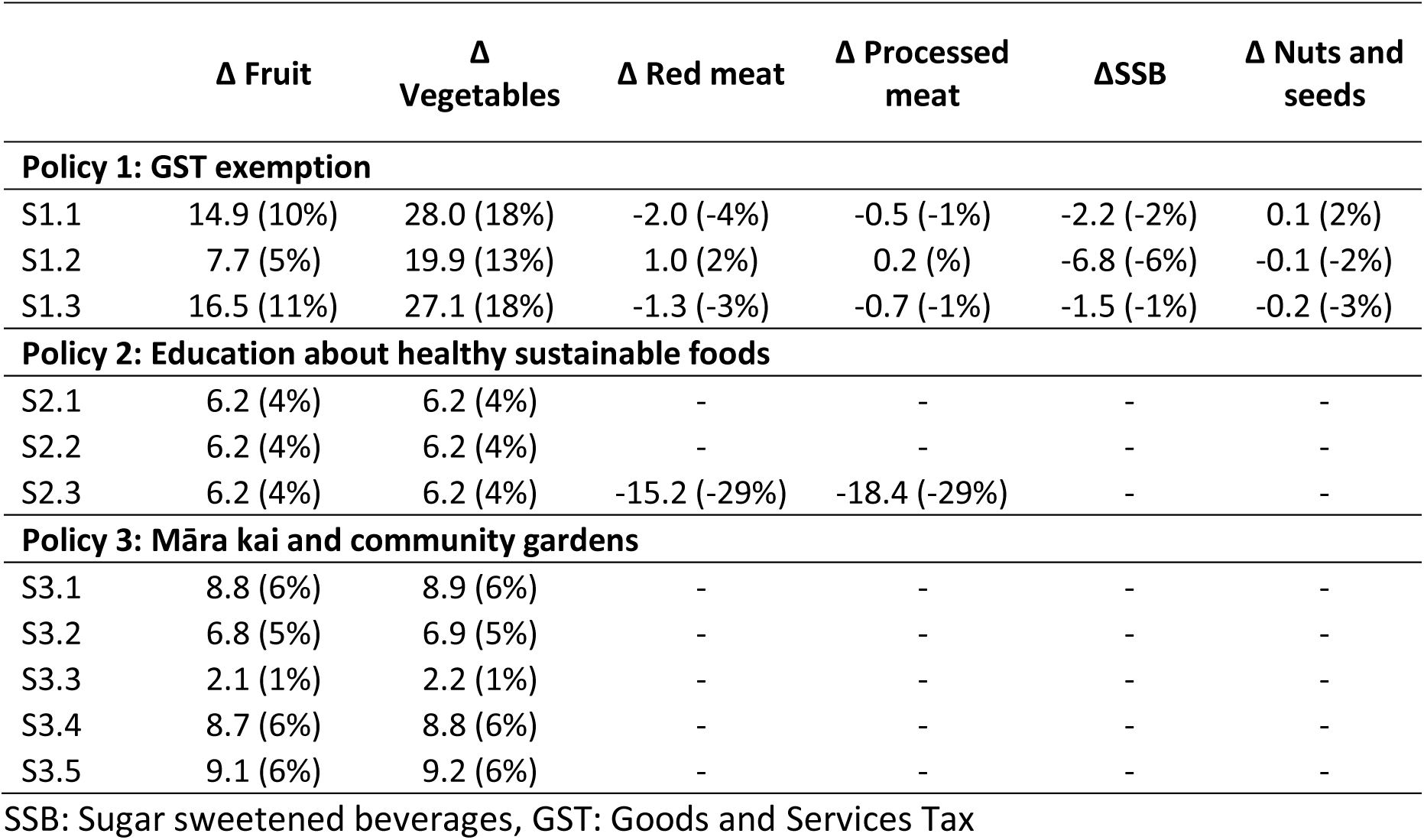
Change in dietary risk factors in grams/day (percentage of baseline intake).

|  | **Δ Fruit** | **Δ Vegetables** | **Δ Red meat** | **Δ Processed meat** | **ΔSSB** | **Δ Nuts and seeds** |
| --- | --- | --- | --- | --- | --- | --- |
| **Policy 1: GST exemption** |  |  |  |  |  |  |
| S1.1 | 14.9 (10%) | 28.0 (18%) | -2.0 (-4%) | -0.5 (-1%) | -2.2 (-2%) | 0.1 (2%) |
| S1.2 | 7.7 (5%) | 19.9 (13%) | 1.0 (2%) | 0.2 (%) | -6.8 (-6%) | -0.1 (-2%) |
| S1.3 | 16.5 (11%) | 27.1 (18%) | -1.3 (-3%) | -0.7 (-1%) | -1.5 (-1%) | -0.2 (-3%) |
| **Policy 2: Education about healthy sustainable foods** |  |  |  |  |  |  |
| S2.1 | 6.2 (4%) | 6.2 (4%) | - | - | - | - |
| S2.2 | 6.2 (4%) | 6.2 (4%) | - | - | - | - |
| S2.3 | 6.2 (4%) | 6.2 (4%) | -15.2 (-29%) | -18.4 (-29%) | - | - |
| **Policy 3: Māra kai and community gardens** |  |  |  |  |  |  |
| S3.1 | 8.8 (6%) | 8.9 (6%) | - | - | - | - |
| S3.2 | 6.8 (5%) | 6.9 (5%) | - | - | - | - |
| S3.3 | 2.1 (1%) | 2.2 (1%) | - | - | - | - |
| S3.4 | 8.7 (6%) | 8.8 (6%) | - | - | - | - |
| S3.5 | 9.1 (6%) | 9.2 (6%) | - | - | - | - |

SSB: Sugar sweetened beverages, GST: Goods and Services Tax

All scenarios for this policy are cost saving to the health system (Table 3). S1.1 generated the largest health impacts (87,340 95% Uncertainty Interval (UI): 72,590 to 104,140) and health system cost savings ($925.0 Million (M), UI: $661.5 to $1220.3M). The next largest health gain (82,260 (68,520 to 97,410)) and health system cost savings ($819.4M ($612.3 to $1067.4M)) are seen in the fruit and vegetable only scenario (S1.3). Undiscounted heath gain and health system cost savings range from 329,570 and $2,104.1M in S1.1, targeting core, sustainable foods, down to 140,270 and $382.0M in S1.2, targeting core foods only (Supplementary material, supplementary table 1).

**Table 3.** Health Adjusted Life Years, health system costs, Incremental Cost-Effectiveness Ratios and age standardised per capita ratio of HALYs between Māori and non-Māori for all scenarios.

| | Lifetime HALYs | Lifetime costs<br>(2011 NZ\$ million) | ICER | Age standardised per capita<br>ratio of HALYs between<br>Māori and non-Māori |
| --- | --- | --- | --- | --- |
| **Policy 1: GST exemption** |  |  |  |  |
| S1.1 | 87,340 (72,590 to 104,140) | \$-925.0 (-661.5 to -1220.3) | Cost saving | 1.4 |
| S1.2 | 38,470 (25,890 to 51,470) | \$-249.8 (151.6 to -577.7) | Cost saving | 2.1 |
| S1.3 | 82,260 (68,520 to 97,410) | \$-819.4 (-612.3 to -1067.4) | Cost saving | 1.3 |
| **Policy 2: Education about healthy sustainable foods** |  |  |  |  |
| S2.1 | 1,170 (930 to 1,430) | \$-9.6 (-6.8 to -12.9) | Cost saving | 1.8 |
| S2.2 | 19,270 (15,550 to 23,700) | \$-156.2 (-117.1 to -206.9) | Cost saving | 1.5 |
| S2.3 | 9,360 (7,170 to 11,500) | \$-165.0 (-122.1 to -217.1) | Cost saving | 2.4 |
| **Policy 3: Māra kai and community gardens** |  |  |  |  |
| S3.1 | 8,040 (4,720 to 11,800) | \$364.3 (267.6 to 464.6) | \$45,300 | 1.4 |
| S3.2 | 6,030 (3,370 to 8,830) | \$317.7 (238.5 to 408.3) | \$52,600 | 1.4 |
| S3.3 | 1,970 (1,170 to 2,880) | \$111.8 (85.2 to 140.0) | \$56,900 | 1.4 |
| S3.4 | 8,030 (4,610 to 11,790) | \$783.3 (606.2 to 968.7) | \$97,500 | 1.3 |
| S3.5 | 8,000 (4,480 to 11,800) | \$0.2 (-47.2 to 40.3) | \$30 | 1.4 |

3 HALY: Health Adjusted Life Years, ICER: Incremental Cost-Effectiveness Ratio

The ratio of age standardised per capita health gain between Māori and non-Māori range from 1.3 (S1.3) to 2.1 (S1.2). For S1.1, health gain for Māori increases by 39% when the equity analysis was applied and the ratio of age standardised, per capita health gain for Māori increased to between 1.9 and 2.9 compared to non-Māori health gain.

### 3.2 Policy 2: Education about healthy sustainable foods

All scenarios have an increase of 6.2 grams (4% of baseline intake) of fruit and vegetables. S2.3 includes a reduction in red and processed meat of 15.2g and 18.3g (both 29% of baseline intake).

S2.1 generates small health impacts (1,170 HALYs, UI: 930 to 1,430)) and health system cost savings ($9.6M, UI: $6.8 to $12.9M). Extending the length of effect from 2 years total (S2.1) to assuming the change in fruit and vegetable intake lasts the cohorts lifetime increased health gain to 19,270 HALYs (UI: 15,550 to 23,700) and health system cost savings to $156.2M (UI: $117.1 to $206.9M). Combining a reduction in red and processed meat with the increase in fruit and vegetable intake (S2.3) give health gains for 9,360 HALYs (UI: 7,170 to 11,500) and health system cost savings of $165.0M (UI: $122.1 to $217.1M). Undiscounted heath gain was 2,170 in S2.1 where fruit and vegetable intake increased for 2 years, 73,910 in S2.2 when the effect lasts the cohorts lifetime, and 17,860 in S2.3, where the policy also decreases red and processed meat intake. Undiscounted health system cost savings follow the same pattern: S2.1: $8.7M, S2.2: $282.1M, S2.3: $225.6M.

The ratio of age standardised per capita health gain between Māori and non-Māori is 1.5, 1.5 and 2.5 for S2.1, S2.2 and S2.3 respectively. In S2.1, health gain for Māori increases by 32% when the equity analysis is applied and the age standardised, per capita health gain for Māori increases from 1.5 times to 2.5 times that of non-Māori.

### 3.3 Policy 3: Māra kai and community gardens

S3.1, with māra kai for all marae and community gardens for all suburbs in NZ (for the subsequent 10 years and a part time (0.5 full time equivalent, FTE) community champion) has the largest increase in average fruit and vegetables (9g of both). This increase is shared with scenarios S3.4 and S3.5 which also cover all marae and suburbs. The scenarios that cover a lower proportion of marae and suburbs have a lower average increase in fruit and vegetable intakes, approximately 7g in S3.2 (All marae and a matched number of suburbs in NZ) and 2g in S3.3 (1/3 of marae and matched number of suburbs in NZ).

The largest health gains were seen for all the scenarios targeting all marae and suburbs (S3.1, S3.4 and S3.5), between 8,000 and 8,040 HALYs. Specifically, for S3.1: 8,040 (UI: 4,720 to 11,800 HALYs) and health system costs of $364.3M (UI: $267.6 to $464.6M). S3.1 was borderline cost-effective with an Incremental Cost Effectiveness Ratio (ICER) of $45,300 per HALY gained (using 2011 $45,000, a governmental willingness-to-pay threshold set to the level of gross domestic product per capita per HALY gained).

Decreasing the number of marae and suburbs receiving this policy (S3.2 and S3.3) reduces both health gain and cost of the policy and the cost-effectiveness ratio increases (moves further away from being cost-effective) to $52,600 (S3.2) and $56,900 (S3.3) per HALY gained. S3.4, where one full time community champion was employed per garden was also not cost-effective with an ICER of $97,500/HALY gained and S3.5, where one community champion looked after 15 gardens, was cost-effective at $30 per HALY gained and only costing $200,000 over the life course of the cohort. Undiscounted heath gain ranged from 4,020 in S3.3 to 16,550 in S3.1, S3.4 and S3.5. Undiscounted health system costs ranged from a cost saving of $3.6M in S3.5 with 1 full time community champion per 15 gardens to health system costs of $778.5M in S3.4, with 1 full time community champion per garden.

The ratio of age standardised per capita health gain between Māori and non-Māori was 1.4 for all scenarios except S3.3 where the ratio was 1.7. In S3.1, health gain for Māori increases by 34% when the equity analysis is applied and the ratio of age standardised, per capita health gain for Māori compared to non-Māori changed from 1.4 to 1.9.

## 4 Discussion

This research modelled the likely health, health equity and health system cost impacts of three policies, which were informed by focus groups and interviews with a range of NZ stakeholders interested in sustainable healthy food systems and consumption. The article provides evidence on the likely health impact if these policies were implemented in NZ, under a range of different assumptions.

Under the first policy, where core, sustainable foods are exempt from GST, moderate health gains and health system cost savings ($925 M) are seen. This modelling does not include the loss of income to the government due to the exemption. Using estimates of 2011 food costs and population numbers we would expect the revenue to the government to decrease by approximately $890 M a year. On balance, we expect all GST exemption scenarios to cost the government money if loss of GST income is considered.

GST exemption affects the whole population and is likely to cause small shifts in population dietary intake over their lifetimes to generate these health impacts. Under the assumption that Māori and non-Māori respond the same to these price changes, larger per capita health gains are generated for Māori suggesting this policy could reduce health inequities between Māori and non-Māori. Additionally, there is evidence that Māori are more sensitive to changes in food prices (Mhurchu, Eyles et al. 2013) which may increase Māori health gain further. Regardless of the mechanism used, these results show that reducing the cost of core foods can have a positive effect on health equity. Other mechanisms to reduce the cost of core food such as increased competition should be explored. The GST exemption policy in Australia is generally attributed to multiple studies finding a ‘healthy’ diet is cheaper than an ‘unhealthy’ diets, however, healthy diets are still found to be unaffordable to marginalised groups such as people on low incomes, and for First Nations people (Dawson, Chung et al. 2025). In NZ, there are equity criticisms of this policy as higher income individuals are likely to receive a larger absolute reduction in their food expenditure. The health benefits of this policy will need to be considered alongside the barriers identified through earlier stages of this project: loss of government revenue; the perceived difficulty associated with defining core and sustainable foods and associated industry pushback; retailers not passing on these savings to the customer, and the stated equity concerns.

There is a lack of evidence on the long-term effect of mass media campaigns on dietary intake so the policy to educate the public on healthy sustainable foods was only modelled to effects on dietary intake in the short term. As such the health gains were relatively small. Under the assumption that dietary intake would change for the lifetime of the cohort health gain increased but only to about a fifth of the GST exemption of core sustainable food policy. Modelling a 46% reduction in red and processed meat intake for a year of the cohort’s lifetime generated 8 times higher health gain than a change in fruit and vegetable intake alone. However, the evidence on the effect of a mass media campaign on red and processed meat intake is weak: participants were only followed up for 1 week (Carfora, Caso et al. 2017). To ensure the relevance and effectiveness of this policy, stakeholders emphasised the importance of a clear definition for ‘sustainable and healthy’ food, suggested implementing this policy alongside other food policies, that the policy be led by the Ministry of Health with cross sector collaboration and that a separate framework for Māori be developed.

The scenarios under the policy to increase māra kai and community gardens varied the number of gardens and the intensity of involvement of community champions. The results show that the most extensive coverage gives the greatest health gain, as would be expected. It is the most expensive (to cover the costs associated with paying community champions) but it is also the scenario which is the closest to being cost-effective. When this champion was part time the policy was borderline cost-effective ($45,300/HALY gained) and when a full-time community champion managed 15 gardens the policy cost $30 per HALY gained, assuming this level of staffing allowed the community gardens to be maintained (mainly by volunteers). Funding for needed resources and strong oversight and leadership to ensure longevity was emphasised by stakeholders when discussing this policy. They envisioned this policy as wider than just community gardens as they could provide spaces to tell Māori stories through kai, opportunities for knowledge transfers, they could act as food hubs and provide connections to local farmers. Barriers such as how to ensure equitable access to gardens and māra kai and how to ensure the longevity of the gardens need to be considered when designing these policies. For this policy in particular there would be wider benefits than the chronic disease reduction modelled. Relevant benefits, illustrated in Figure 1, include strengthening local food networks, improved food security and food system resilience alongside benefits to social cohesion and wellbeing.

Although all policies are either cost saving or could be designed to be cost-effective, reductions to health system expenditure occur over a long time period (the lifetime of the 2011 cohort). For education about healthy sustainable foods and māra kai and community gardens these require up front expenditure and GST exemption will lose government revenue. Governments need long-term vision to value the benefits of such policies. High and rising rates of diet related disease need to be addressed. Implementing a variety of policies with both short- and long-term impacts on the food environment and food choices is key to reducing these disease rates. They should also consider the additional benefits of these policies alongside reductions in chronic disease (e.g., impacts on food security, social cohesion, wellbeing and food system resilience).

All modelled policies generated more age standardised per capita health gains for Māori than non-Māori with policy 2, education about healthy sustainable foods, having the biggest difference in per capita health gain between Māori and non-Māori. There are a number of assumptions that would need to be met for these greater health gains to be realised, especially for policy 2 which requires careful design and targeting to ensure positive dietary change is either the same or greater for Māori. All policies should be designed by and with Māori to meet the assumptions around equal uptake and effectiveness for Māori.

There is an argument that undiscounted modelling results should be prioritised as disease developing in middle to late life of those that are children now should be valued the same as health gain generated in current adults. The undiscounted health gain is between 1.9 and 3.8 times greater than the health gain discounted at 3% but the relative health gain between the different policies and scenarios does not change.

The modelled policies were selected by stakeholders due to their likely ability to improve NZ diets, Māori health, and the environment as well as being feasible and acceptable. We were not, however, able to model the impact on GHG emissions of all policies due to a lack of data on their likely impact on emissions. Policy 2 and 3 increased fruit and vegetable intake but there was no evidence of what foods this increase might displace (if any) in the diets. We therefore cannot comment on the climate impact of these policies, despite them being selected and designed to be environmentally sustainable.

### 4.1 Policy implications

Evidence of policy effectiveness, the length of effect and policy uptake were based on meta-analyses (Afshin, Penalvo et al. 2015), if available, or the best, most relevant international evidence available (Dixon, Borland et al. 1998, Alaimo, Packnett et al. 2008, Carfora, Caso et al. 2017, Morley, Niven et al. 2019, Morley, Nuss et al. 2022), except for the GST policy which was based on NZ price elasticities (Mhurchu, Eyles et al. 2015). These intervention parameters were applied to NZ specific demographic, diet and disease data (Cleghorn, Blakely et al. 2017). Countries with comparable populations and disease rates could use these results to inform their own policy design and especially to compare the relative benefits of policies. The results from the current analyses can be compared with other dietary policies published using the same dietary model including a range of food and beverage taxes (Blakely, Cleghorn et al. 2020, Grout, Mizdrak et al. 2022), a food tax based on GHG emissions (Cleghorn, Mulder et al. 2022) and interventions targeting individual behavior change (Cleghorn, Wilson et al. 2020, Jones, Grout et al. 2022).

To understand and model these policies more accurately, further research on their effectiveness, their long-term effects, and their impacts on total dietary intake are needed. Additionally, further research is needed into the effect on dietary intake in the medium and long term of a mass media campaign specifically on sustainable healthy dietary intake. This research has shown that the three modelled policies have the potential to improve population health and reduce ethnic health inequities in NZ. Due to the complementary nature of these policies, they could be implemented simultaneously to maximise impact. Based on the results of the current study, we recommend GST exemption for core, sustainable foods, alongside support to marae and communities to establish mara kai and community gardens across the country. These policies could be supported by a comprehensive and well-maintained mass media campaign to promote these policies, increase their social acceptance, and encourage healthy sustainable eating. Careful design involving relevant stakeholders including Māori communities are needed to ensure health benefits are realised and the wider social benefits should also be considered. Results can be used to better understand the impact these policies could have in other high-income countries and their relative benefits.

## Data Availability

Data used in the present study are not available due to data use agreements

## Acknowledgements

The authors thank those who took part in the qualitative research which informed the policy design used in this paper. They thank those who contributed to models used in this work (Tony Blakely, Nick Wilson, Linda Cobiac, Anja Mizdrak, Giorgi Kvizhinadze, Nhung Nghiem). The authors thank the 4,721 New Zealanders who participated in the 2008/09 New Zealand Adult Nutrition Survey. The New Zealand Ministry of Health funded the 2008/09 New Zealand Adult Nutrition Survey. Access to the data used in this study was provided by Statistics New Zealand under conditions designed to keep individual information secure in accordance with requirements of the Statistics Act 1975. Data interpretation do not necessarily represent an official view of Statistics New Zealand. Figure 1 was illustrated by Yasmine El Orfi (www.yasmineelorfi.com).

## Author contributions

C Cleghorn led the conceptualization of the research; analysis (modelling); funding acquisition; methodology; investigation; project administration; visualization; writing of the original draft, reviewing and editing the manuscript. C McKerchar was involved in the methodology; investigation; and contributed to the original draft of the manuscript. B Kidd was involved in the analysis (modelling); methodology; investigation; project administration and contributed to the original draft of the manuscript. C Ni Mhurchu was involved in the conceptualization of the research; funding acquisition; methodology and contributed to the original draft of the manuscript.

