## Supplementary material for "Modelled health impacts of three stakeholder-selected policies to support healthy and environmentally sustainable population diets"

**Supplementary material: Glossary of Māori words from Te Aka Māori dictionary (https://maoridictionary.co.nz)**

Iwi: extended kinship group, tribe, nation, people, nationality, race - often refers to a large group of people descended from a common ancestor and associated with a distinct territory.

Kai: food, meal.

Mana Motuhake: separate identity, autonomy, self-government, self-determination, independence, sovereignty, authority - mana through self-determination and control over one's own destiny.

Māori: Indigenous New Zealander.

Māra: garden.

Māra kai: gardening for food.

Marae: Courtyard - the open area in front of the meeting house, where formal greetings and discussions take place. Often also used to include the complex of buildings around the marae.

Te Tiriti o Waitangi: The Treaty of Waitangi.

### Supplementary table 1. Health adjusted life years and health system costs for all scenarios

| | Non-Māori | Māori | Māori | Ethnic groups combined | |
|---|---|---|---|---|---|
| | HALYs | HALYs | equity analysis HALYs* | HALYs | Costs (2011 NZ$ million) |
| **Policy 1: GST exemption for core and sustainable foods, time horizon: lifetime of cohort** | | | | | |
| **S1.1** | **Core sustainable foods** | | | | |
| Sex groups combined | 70,620 (58,430 to 84,150) | 16,730 (13,460 to 20,210) | 23,210 (19,160 to 27,670) | 87,340 (72,590 to 104,140) | $-925. (-661.5 to -1220.3) |
| Men | 37,060 | 8,760 | 12,040 | 45,820 | |
| Women | 33,560 | 7,970 | 11,170 | 41,530 | |
| Per capita* | 18.9 (18.4) | 24.8 (25.4) | 34.4 (35.5) | 19.8 | -$ 210.0 |
| 0% discounting | 262,310 | 67,260 | 101,630 | 329,570 | -$ 2,104.1 |
| **S1.2** | **Core foods only** | | | | |
| Sex groups combined | 28,500 (20,090 to 37,450) | 9,960 (4,230 to 15,130) | 13,580 (8,850 to 18,540) | 38,470 (25,890 to 51,470) | $-249.8 (151.6 to -577.7) |
| Men | 15,230 | 6,220 | 8,270 | 21,450 | |
| Women | 13,280 | 3,740 | 5,310 | 17,020 | |
| Per capita* | 7.6 (7.2) | 14.8 (15.0) | 20.1 (20.6) | 8.7 | -$ 56.7 |
| 0% discounting | 102,820 | 37,450 | 56,600 | 140,270 | -$ 382.0 |
| **S1.3** | **Fruit and vegetables only** | | | | |
| Sex groups combined | 66,990 (55,760 to 79,320) | 15,270 (12,780 to 18,140) | 21,280 (17,830 to 25,180) | 82,260 (68,520 to 97,410) | $-819.4 (-612.3 to -1067.4) |
| Men | 35,080 | 7,270 | 10,070 | 42,350 | |
| Women | 31,910 | 8,000 | 11,200 | 39,910 | |
| Per capita* | 18.0 (17.4) | 22.7 (23.2) | 31.6 (32.5) | 18.7 | -$ 186.0 |
| 0% discounting | 250,430 | 62,290 | 94,280 | 312,720 | -$ 1,855.2 |
| **Policy 2: Education about healthy sustainable foods** | | | | | |
| **S2.1** | **Assume the effect on fruit and vegetable intake is maintained for two years** | | | | |
| Sex groups combined | 980 (770 to 1,220) | 190 (150 to 230) | 250 (200 to 310) | 1,170 (930 to 1,430) | $-9.6 (-6.8 to -12.9) |
| Men | 540 | 90 | 120 | 630 | |

| | | | | | |
|---|---|---|---|---|---|
| Women | 440 | 100 | 130 | 540 | |
| Per capita* | 0.3 (0.2) | 0.3 (0.3) | 0.4 (0.5) | 0.3 | -$ 2.2 |
| 0% discounting | 1,810 | 360 | 510 | 2,170 | $-8.7 |
| **S2.2** | **Assume the effect on fruit and vegetable intake is maintained for the lifetime of the cohort** | | | | |
| Sex groups combined | 15,480 (12,290 to 19,160) | 3,790 (2,960 to 4,720) | 5,410 (4,220 to 6,700) | 19,270 (15,550 to 23,700) | $-156.2 (-117.1 to -206.9) |
| Men | 8,230 | 1,800 | 2,570 | 10,030 | |
| Women | 7,260 | 1,990 | 2,840 | 9,240 | |
| Per capita* | 4.2 (4.0) | 5.6 (5.8) | 8.0 (8.3) | 4.4 | -$ 35.5 |
| 0% discounting | 58,100 | 15,820 | 24,460 | 73,910 | $-282.1 |
| **S2.3** | **Scenario including a reduction of red and processed meat by 46%** | | | | |
| Sex groups combined | 7,280 (5,570 to 9,020) | 2,080 (1,610 to 2,540) | 2,590 (2,010 to 3,230) | 9,360 (7,170 to 11,500) | $-165. (-122.1 to -217.1) |
| Men | 4,190 | 1,110 | 1,390 | 5,290 | |
| Women | 3,090 | 980 | 1,210 | 4,070 | |
| Per capita* | 2.0 (1.5) | 3.1 (3.7) | 3.8 (4.6) | 2.1 | -$ 37.5 |
| 0% discounting | 13,950 | 3,910 | 5,260 | 17,860 | $-225.6 |
| **Policy 3: Mara kai and community gardens, time horizon: 10-years** | | | | | |
| **S3.1** | **All marae and suburbs in NZ (0.5 FTE community champion/garden)** | | | | |
| Sex groups combined | 6,940 (3,600 to 10,560) | 1,100 (540 to 1,700) | 1,470 (760 to 2,280) | 8,040 (4,720 to 11,800) | $364.3 (464.6 to 267.6) |
| Men | 3,790 | 520 | 700 | 4,310 | |
| Women | 3,140 | 580 | 770 | 3,720 | |
| Per capita* | 1.9 (1.4) | 1.6 (1.9) | 2.2 (2.6) | 1.8 | $ 82.7 |
| 0% discounting | 14,210 | 2,330 | 3,300 | 16,550 | $ 359.6 |
| **S3.2** | **All marae and matched number of suburbs in NZ (0.5 FTE community champion/garden)** | | | | |
| Sex groups combined | 5,180 (2,640 to 7,870) | 860 (440 to 1,330) | 1,120 (530 to 1,750) | 6,030 (3,370 to 8,830) | $317.7 (408.3 to 238.5) |
| Men | 2,830 | 410 | 530 | 3,240 | |
| Women | 2,340 | 450 | 590 | 2,800 | |
| Per capita* | 1.4 (1.1) | 1.3 (1.5) | 1.7 (2.0) | 1.4 | $ 72.1 |
| 0% discounting | 10,550 | 1,820 | 2,580 | 12,380 | $ 316.2 |
| **S3.3** | **1/3 of Marae and matched number of suburbs in NZ (0.5 FTE community champion/garden)** | | | | |
| Sex groups combined | 1,690 (910 to 2,580) | 280 (140 to 440) | 370 (180 to 580) | 1,970 (1,170 to 2,880) | $111.8 (140. to 85.2) |

| | | | | | |
|---|---|---|---|---|---|
| Men | 920 | 130 | 180 | 1,050 | |
| Women | 760 | 150 | 200 | 910 | |
| Per capita* | 0.5 (0.3) | 0.4 (0.5) | 0.6 (0.7) | 0.4 | $ 25.4 |
| 0% discounting | 3,430 | 590 | 840 | 4,020 | $ 111.2 |
| **S3.4** | **All marae and suburbs in NZ (1 FTE community champion/garden)** | | | | |
| Sex groups combined | 6,950 (3,610 to 10,490) | 1,090 (530 to 1,730) | 1,450 (710 to 2,280) | 8,030 (4,610 to 11,790) | $783.3 (968.7 to 606.2) |
| Men | 3,810 | 510 | 690 | 4,320 | |
| Women | 3,140 | 570 | 760 | 3,710 | |
| Per capita* | 1.9 (1.4) | 1.6 (1.9) | 2.1 (2.6) | 1.8 | $ 177.8 |
| 0% discounting | 14,210 | 2,330 | 3,300 | 16,550 | $ 778.5 |
| **S3.5** | **All marae and suburbs in NZ (1 FTE community champion/15 gardens)** | | | | |
| Sex groups combined | 6,910 (3,440 to 10,670) | 1,090 (530 to 1,720) | 1,450 (740 to 2,240) | 8,000 (4,480 to 11,800) | $.2 (40.3 to -47.2) |
| Men | 3,790 | 520 | 680 | 4,300 | |
| Women | 3,120 | 570 | 760 | 3,690 | |
| Per capita* | 1.9 (1.4) | 1.6 (1.9) | 2.1 (2.6) | 1.8 | $ - |
| 0% discounting | 14,210 | 2,330 | 3,300 | 16,550 | -$ 3.6 |

*Per capita (HALYs /1000 people and $). FTE: Full-time equivalent.
